# Forecasting virus outbreaks with social media data via neural ordinary differential equations

**DOI:** 10.1101/2021.01.27.21250642

**Authors:** Matías Núñez, Nadia L. Barreiro, Rafael A. Barrio, Christopher Rackauckas

## Abstract

In the midst of the covid-19 pandemic, social media data collected in real time has the potential of being an early indicator of a new epidemic wave. This possibility is explored here by using a neural ordinary differential equation (neural ODE) that is trained to predict virus outbreaks for a geographic region. It learns from multivariate time series of signals obtained from a novel set of massive online surveys about COVID-19 symptoms. Once trained, the neural ODE is able to capture the dynamics of the interlinked local signals and accurately predict the number of new infections up to two months in advance. Moreover, it can estimate the future effects of changes in the number of infected at a given time, which can be associated with the flow of people entering or leaving a given region or, for instance, with a local vaccination campaign. This work gives compelling preliminary evidence for the predictive power of widely distributed social media surveys for public health application

## I. INTRODUCTION

During a pandemic, the ability to identify and forecast local virus outbreaks is key in order for health officials to take appropriate action. However, the intrinsic parameters that represent the biological properties of the virus, used by the predicting models, can only be estimated *once* the pandemic has occurred. While a pandemic is happening, parameter estimation carries a large degree of uncertainty, meaning that the first principles models that use them inherit this uncertainty in their predictions. An epidemiologist recently pointed out in the New York Times^1^: “*You tell me what numbers to put in my equations, and I’ll give you the answer* … *But you can’t tell me the numbers, because nobody knows them*…*”*, a statement that illustrates the difficulties that currently exist in predicting new infections during a pandemic.

A large amount of data is being generated, directly or indirectly, related to the spread of the virus, on various spatial and temporal scales. Sources include social media, mobile phone GPS, and mobile fitness devices, with other tools such as contact tracing or contact simulations^2^. It is clear that it is necessary to include this information in the first principles models or combinations with data-driven models^3,4^ through a learning process or optimization.

In this work we investigate the predictive power of data-driven models using survey data related to COVID-19 captured through Facebook. Signals in function of time are extracted from the data for a given geographic region. They are obtained after processing the survey by extracting numerical indicators (signals) from questions related to people’s symptoms, infections among their social circle, visits to the hospital, quantity of online searches about COVID-19, average time away from home and more related questions. For instance, a person response about how many of his friends tested COVID positive will be somehow correlated with the new infected cases in his region.

There is no clear first principles model for connecting the factors of the survey with the COVID-19 statistics. However, it is reasonable to expect that the local variation in time of the survey responses for a region is correlated with new virus infections in that region. Moreover, these signals have the potential of being early indicators^5^, as they are not subject to intrinsic delays related to the officially reported variables, local policies or testing capacities.

In order to learn this relation, a neural ordinary differential equation (neural ODE)^6^ was used to parameterize the signals rate of change. This object uses a parameterized universal approximator in order to represent all possible phase space dynamics with a finite set of parameters that can be learned on the training data. In this work the neural ODE is trained on these potential early indicators and is able to predict virus outbreaks even two months in advance. Moreover, once trained, these phase space methods allow for forecasting possible future scenarios.

Section I A details the surveys and signals as well as arguments that support the idea that these signals could be used as early indicators. Section I B gives a brief description of neural ODEs and how they are used in this work. Section II details the specific methods for incorporating the data into the neural ODEs and Section III demonstrates the predictive power of the neural ODEs when used in this capacity. We end by discussing the ramifications of using these machine learning methods and data in the context of health care statistics.

### A. COVID-19 Symptom Surveys through Facebook

Since April 2020 universities and public health officials, in collaboration with Facebook, have been conducting a massive daily survey to monitor the spread and impact of the COVID-19 pandemic in the United States. The survey^7^ is an ongoing operation that is advertised through Facebook’s platform and is taken by nearly 55,000 people every day. Respondents provide information about COVID-related symptoms, contacts, prior medical conditions, risk factors, mental health, demographics and the economic effects of the pandemic. The information allows researchers to examine county-level trends across the US. Around 16 million responses have been collected so far.

The survey has four sections and it contains 35 questions. The first section gathers information about a set of symptoms used to define a condition called COVID-like illness (CLI), defined as fever of at least 100 °F, along with shortness of breath, difficulty breathing or a cough^8^. Two key quantities are estimated with this information, for a given location and day:

1. the percentage of people with CLI,
2. the percentage of people who know someone in their local community with CLI illness (CLI-in-community).

The second section gathers more details on testing, symptoms, and medical-seeking behavior. The third section collects info on contacts and risk factors, and the fourth section on demographics. A sample of the exact questions asked can be found in the Supplementary Materials. Numerical indicators (signals) are extracted from the set of questions^7^ that results in a set of time series (one for each indicator), for a given location. The aggregated data is publicly available on the Delphi Group websites^9^,^10^.

#### 1. Surveys as early indicators

A person normally experiences viral symptoms before seeking a COVID-19 test or medical care. Therefore, data related to how many people are self-reporting CLI symptoms in a given location could potentially give an early indicator of COVID activity in that location. Moreover, the data is not subject to reporting delays, unlike formal testing metrics of confirmed daily COVID-19 cases, which are also affected by issues such as testing policy and capacity.

An analysis that provides evidence that the survey-based CLI signals can be early indicators of COVID activity is provided in the Delphi Group^11^ where it is shown that the “CLI-in-community” signal rises alongside confirmed COVID-19 cases. Indeed, more people report that others are sick in their community at times when COVID-19 tests confirm more cases. Interestingly, the indicator begins to rise steeply days ***before*** COVID-19 cases begin their steep ascent. This analysis is an informal way of looking at the recall of the indicator and opens up the possibility of using these kind of noisy and indirect signals as early indicators of new cases.

Although the survey cannot be used to draw definite conclusions about the true prevalence of coronavirus disease in the studied region, ***changes in self-reported symptoms over time*** could still be a meaningful reflection of the changes in coronavirus infections over time and therefore could help predicting changes in the number of new infected cases that will happen some days into the future.

### B. Models: first principles and data driven

The use of these signals for the prediction of new cases could be done by means of a model that relates the rate of variation of the different indicators to the model’s state variables. However, unlike the case of common epidemiological models, the deduction of a quantitative expression that relates the new cases as a function of the different signals extracted from the surveys is far from obvious. Even if the relations were found and the model described for example as an ordinary differential equation system, it would surely have unknown parameters involved and would be subject to uncertainties. Taking advantage of the amount of data and indicators obtained from the surveys, a data driven approach would be a logical alternative to obtain a model for predicting the new infected cases for a geographical region.

Therefore, for a specific region, we define a vector with a suitable set of indicators / variables as components (among them the number of new cases) and define the model via a function that approximates the temporal evolution of this vector. With such a function, the number of new cases is expressed as function of time and thus a prediction could be made. For example, in the case of the classic SIR compartment model^12^, the vector components are the variables number of suspectible individuals (S), the number of infected individuals (I) and the number of recovered individuals (R) and their temporal variation expressed with via an ordinary differential equation based on intuition and qualitative knowledge of the dynamics of contagions. But for the case of the vector formed with the surveys indicators as components, since it is not clear how to define the relationship between them from first principles, we cannot directly define such a model.

#### 1. Neural ordinary differential equations

Given the lack of a known function form, we turn to the neural ODE^6^ as a way to directly derive the differential equation from data. A Neural ODE is a neural network parametrization of an ordinary differential equation which allows for learning the dynamics of any possible dynamical system due to the universal approximation theorem^13,14^ (assuming a sufficiently large neural network). In particular, we represent our dynamical system via:

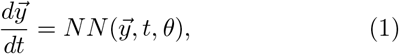

where *NN* is a neural network given by weights *θ*. This neural network has an explicit *t* dependence since it is parameterized based on the time-dependent input signals from the data. The goal is to learn the underlying dynamics of change. The “forward pass” through a neural ODE is equivalent to solving an initial value problem where *y*(*t*_0_) is the input features and we replace hand-crafted equations with a neural network. A single forward pass gives us an entire trajectory. In contrast to other architectures used for time series like residual neural networks (RNNs)^15^, this model is continuous in time, allowing for incorporating non-uniform data and predictions.

The parameters of the neural ODE are learned from the data as diagrammed in Figure 1. The learning process is performed by minimizing the following loss function

**FIG. 1.**
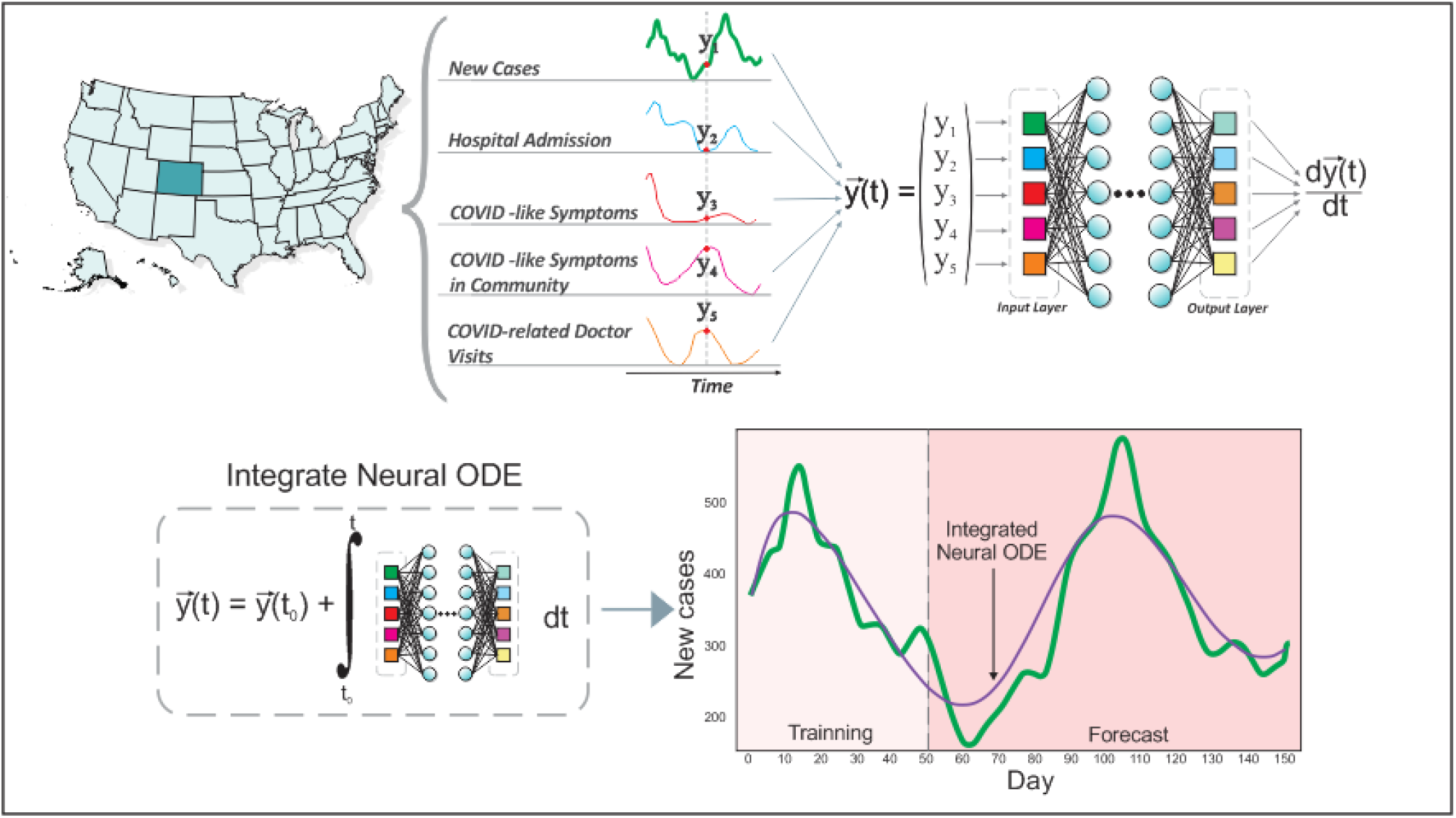
The Neural ODE is trained with a set of signals/variables (shown above) extracted from the online surveys. The trained neuronal net is able to capture the dynamics of the temporal variation of the signals by finding the ordinary differential equation that best describes the data. The learnt solution, obtained by the temporal integration of the neural ODE, is shown below against the reported data for the new infected cases in CO (correspondent to the signal *Y*_1_). The solution spans the training interval and the prediction.

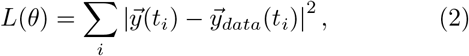

with respect to the networks parameters *θ*. Here, 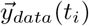 represents the multivariate time series as a vector whose components are the values from the chosen set of signals at time *t*_*i*_ while *y*(*t*_*i*_) is given by the numerical solution to Equation 1. Minimization is performed by gradient-based local methods, specifically ADAM^16^. Thus in order to perform the minimization the gradient of the loss function with respect to all parameters *θ* must be computed. Given the large Lipshitz constants seen due to rapid changes during the onset of the growth, the adjoint technique of the original neural ODE publication is potentially unstable on the case of interest^17–19^, and thus we opted for stabilized techniques which avoid reverse solving^20,21^.

## II. METHODS

The raw signals for each USA State were downloaded using the Delphi Group API^7,10^. A smoothing was performed via a cubic spline interpolation for all the signals/indicators^22^ time series. The 7-day averaged of reported new confirmed COVID-19 cases was used as the main indicator of interest^23^ for accounting for the new cases. We chose the following set of variables as components in order to build the state vector *y*(*t*) for each location:

1. New daily cases (7 day averaged),
2. Hospital Admission,
3. COVID-Like Symptoms,
4. COVID-Like Symptoms in Community,
5. COVID-Related Doctor Visits

The resultant multivariate time series were split into two sets, a training set and a validation set. The training set was used to update the weights *θ* in the network while the validation set was used for monitoring over-fitting and training generalization. Training was performed using a mini-batched^24^ form of multiple shooting^25–27^ which involved computing the loss between intervals of data points. Specifically, a data point was randomly selected to be the initial condition and the neural ODE was solved from the point *y*_*i*_ at time *t*_*i*_ to time *t*_*i*+1_ with the Tsit5 method^28^ using the DifferentialEquations.jl implementation^29^ to get the prediction for the next point *y*(*t*_*i*+1_). This was then compared to the true data point 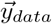. The loss was calculated as the mean squared error (MSE) between the true point 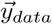 and the predicted point *y*(*t*) (see eq. 2). Backpropogation was performed using the adjoint implementations of the DiffEqFlux.jl library^3^.

The neural networks used for parameterizing the ODE consisted of four interconnected layers with 64, 32, 16 and 8 neurons each and swish activation functions^30^.

Once the weights of neuronal net (*θ*) are found, the network defines the rate for the temporal evolution of the state variables (See eq. 1). Note that such an equation can be solved beyond the training interval to assess its ability to accurate forecast. It is expected that this prediction will deteriorate as it moves further in phase space from the training data, nonetheless we will show that there will be a highly predictive time frame.

### A. Software

Python library Pandas^31^ was used for pre-processing the data^22^. The set of tools available in the Julia library The following open source software tools were used for this work: Pandas library ^31^ for part of the data pre processing, matplotlib^32^ for plotting, and Inkspace for making figures^33^, DiffEqFlux^3,34^ for training the neural ODEs, and the DifferentialEquations.jl solvers^29^ for solving the differential equations.

## III. RESULTS AND DISCUSSION

Figure 2 shows the results of the trained neural ODE and its projection for the state of Ohio. One hundred days of data were used for training. The prediction of the neural ODE follows the thread of the new reported cases fifty days in the future (see interval after the vertical dotted line). Meanwhile, Figure 3 shows the case of the state of Maine where the neural ODE learns to qualitatively extrapolate the new infected cases for 40-50 days also using data from the previous 100 days. The dynamic of the outbreak until day 100 (last training day), looking at the new cases alone, shows a diminish of contagious nonetheless the neural ODE is able to predict an increase on the new cases for the following fifty days.

**FIG. 2.**
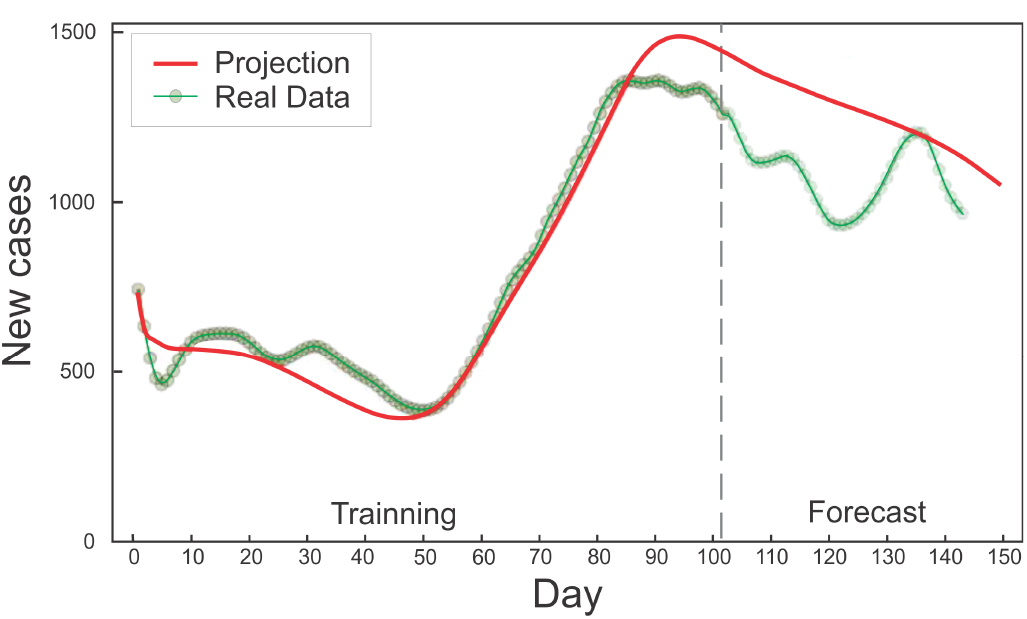
Neural ODE model component for the new cases in the state of Ohio. New reported cases with dots and neural ODE solution with solid line. The vertical dotted line delimits the data set used to train the neuronal network, from the test set. The variables used for this forecast are new cases, Hospital Admission, COVID-Like Symptoms, COVID-Like Symptoms in Community and COVID-Related DoctorVisits^7^.

**FIG. 3.**
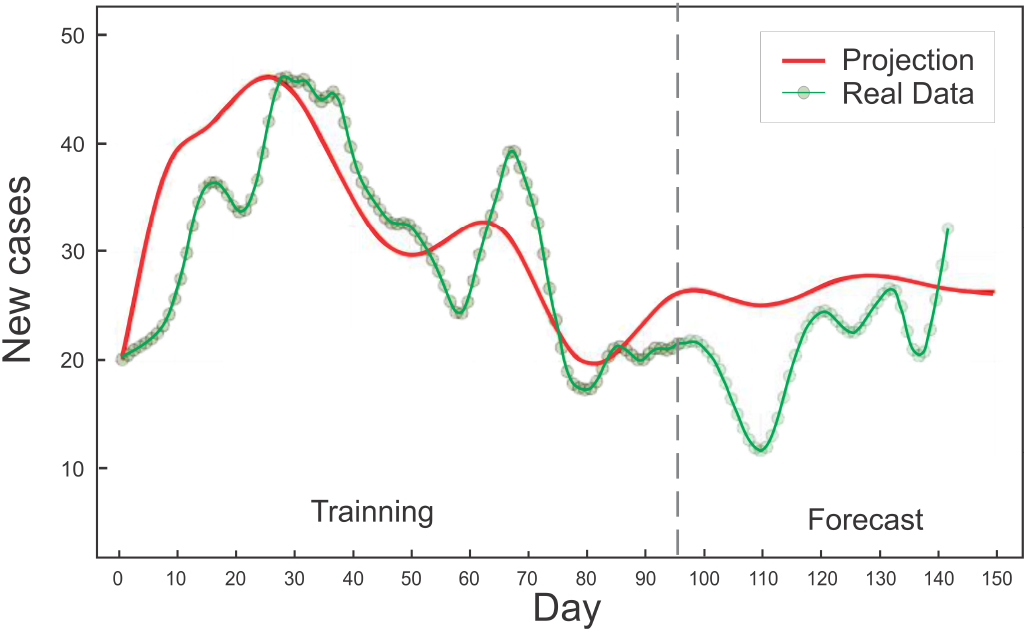
Prediction for the new infected cases in the state of ME (Maine), particular case where the dynamic is not easily describable by a first principles model, while the neural ODE is able to learn the dynamics and predict an increase of cases for the following fifty days. This increase correlates nicely with the recorded cases not used for the learning process (test dots on the right of the vertical line).

Figure 4 demonstrates that in the state of the neural ode is trained with only 50 days of data but is able to extrapolate the outbreak dynamics for the next sixty days, predicting the uprising in the number of cases during the next 15 days after the last training day. Moreover, is able to forecast the day of the next peak of cases and the following decrease of cases.

**FIG. 4.**
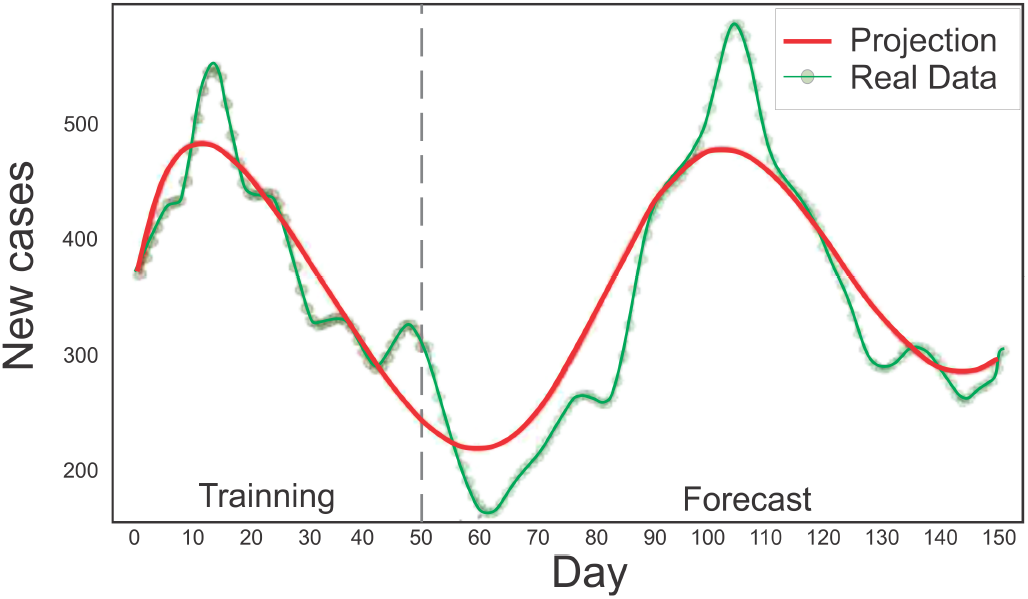
The training data set used here (CO state) finished before another uprising of infected cases. Nonetheless the neural ODE is able to detect the outbreak and even accurately pinpoint almost 60 days later the date of the outbreak peak.

The model (the neural ODE), once it learnt the dynamics from the local signals, is capable of obtaining a prediction of new contagions, but also allows studying possible future scenarios in the event of signal disturbances. For example, if there is an abrupt change in the number of new cases in a given time, the model can predict the effect on the forecast without having to be re-trained. This translates into changing the initial condition in the integration and can be used to estimate forecasting errors due to uncertainty in the signal related to the current number of cases. Figure 5 demonstrates a projection which includes such uncertainty in current epidemic statistics.

**FIG. 5.**
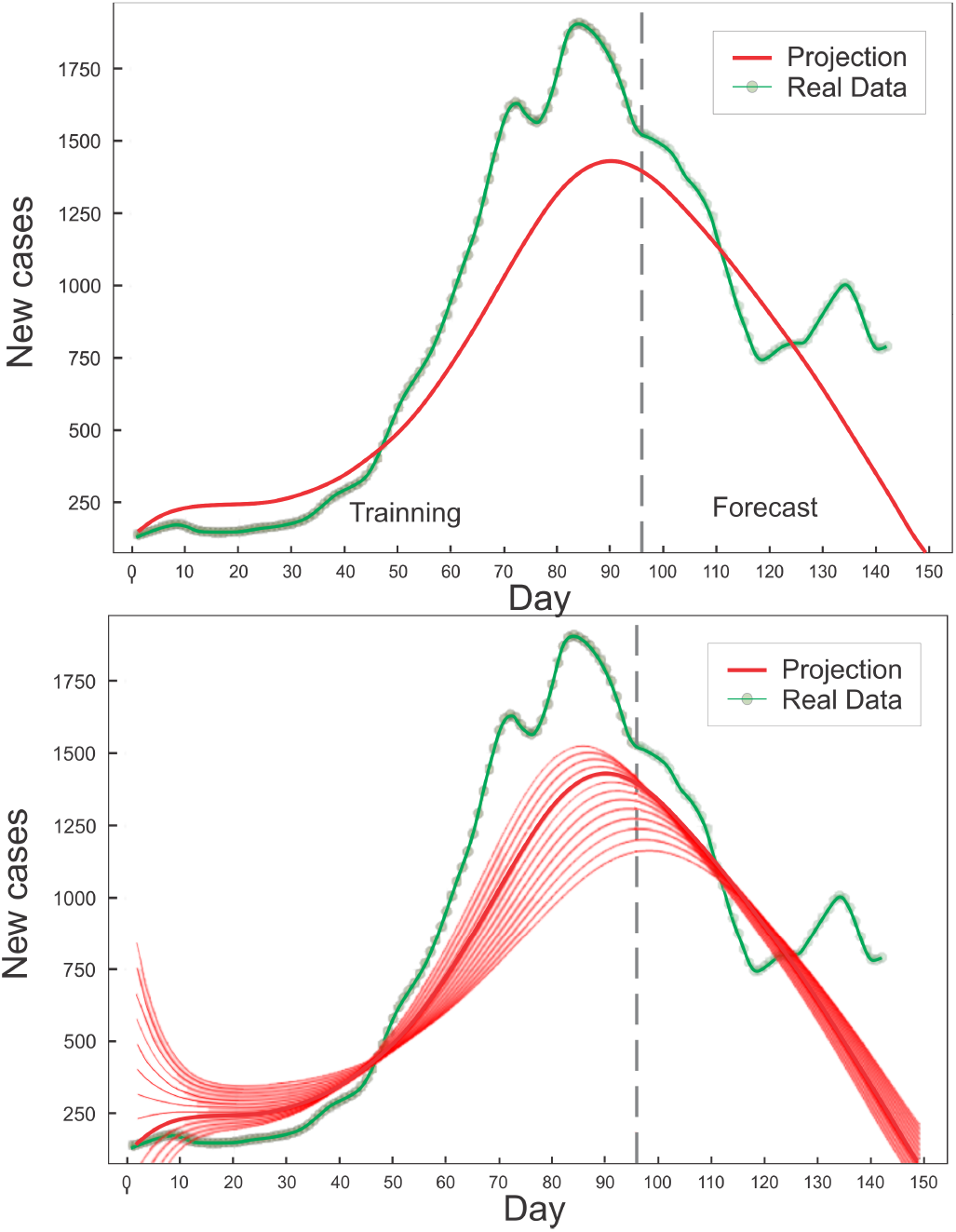
Once the dynamics is learnt by the neural ODE, its solution can predict the projection for different number of initial cases. Above: new cases for the state of South Carolina (dots), and the solution given by the trained neural ODE(thick line). Below: different solutions calculated by the neural ODE for different number of cases on the first day. This ability to estimate the effect of signal perturbation can be useful for estimating the error on the forecast, and also for inferring the effect of changes in the number of cases.

Moreover, a change in the number of infected can be for instance due to the flow of people traveling in or out of the location, which would result in an overall perturbation in the number of infected. Figure 6 showcases the differing levels of the predicted peak with respect to different choices for the amount of migration. The change and its consequences can be estimated by looking at the perturbed solution. This opens the possibility, for locations with strict closed borders to be able to predict the effect of the flow of people on the curve of infected. Moreover, the effects of vaccination in the region could also be estimated in this way.

**FIG. 6.**
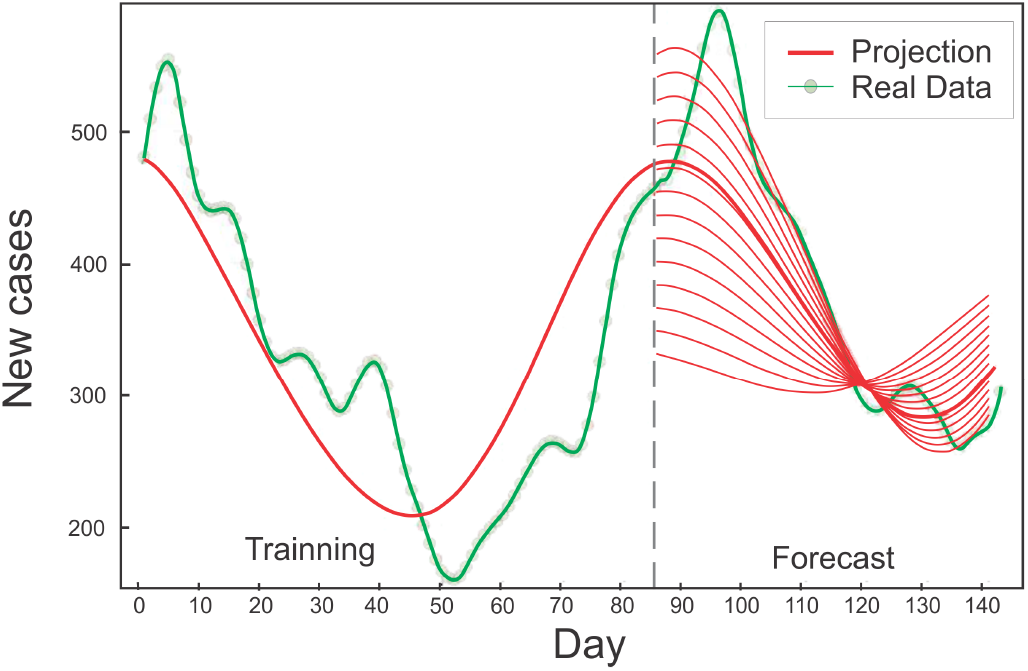
Once trained with the local signals (Colorado state), the neural ODE can extrapolate different possible scenarios due to changes in the number of local active case. This change could happen due to the flow of people traveling in or out of the location or by a local vaccination campaign. The neural ODE solution is shown with a thick line, while the reported data 7 day average of new cases with dots. The test data starts after the vertical dotted line where different changes on the number of infected are defined whit the neural ode forecast shown for each case (thin line).

## IV. CONCLUSIONS AND FUTURE WORK

Using multivariate time series associated with a geographic region, obtained by quantifying indicators from massive online surveys on COVID symptoms offered through the Facebook platform, we show how a neural ODE is able to learn the dynamics that connect these variables and detect virus outbreaks in the region. Analyzing data from different US states, we show that the neural ODE is capable of predicting up to sixty days into the future in different kinds of virus spreading environments.

We show that once it learnt the dynamics of the local signals/variables, the neural ODE is capable of not only forecasting new contagious in the region but also analyzing possible future scenarios in the case of abrupt changes in the number of infected in a given day, for example *due to transit of people to or from the analyzed region or to a vaccination campaign*. This opens the possibility for locations with strict closed borders, to be able to predict the effect of the flow of people on the curve of infected and thus design policies accordingly in a controlled way^35^. Likewise, it could be useful for the design of the strategies of vaccination campaigns.

The neural ODE was trained with data from a single location (state), however it would be expected that the dynamics that connects the local signals in one region will have similar attributes in another. Thus it would be interesting to explore different training schemes, where the model also learns from other regions. Including graphical models into the neural ODE, possibly via graph neural networks, is a compelling avenue for future research.

It is possible to combine this class of data-driven models with first principles models such as compartment models with a scientific machine learning approach^21^. If one region is describable with an analytical model and another is not, but signals can be extracted from it, a hybrid model for the combined region can be designed, with first principles for the first region and with a trained neural ODE for the second.

This work represents a first step, a *proof of concept*. It is necessary to explore different signals and combinations, and compare its generalization capabilities. Accurate usage of the uncertainty quantification also needs much more research before being deployed in public health scenarios. Still, these results showcase promising results for future real-time forecasting from predictive social media data.

## Data Availability

it is  available.

## CODE AVAILABILITY STATEMENT

The code supporting the current study have not been deposited in a public repository because is part of an on-going research. It will be available from the Lead Contact on request.

## AUTHOR CONTRIBUTIONS

Conceptualization - M.N. Methodology - M.N., C.R. Software - M.N., C.R. Writing – Original Draft M.N. Writing – Review Editing M.N., R.A.B.,N.L.B., C.R. Visualization - M.N., N.L.B. Supervision - M.N., R.A.B. Funding Acquisition - M.N., R.A.B., C.R.

## ACKNOWLEDGMENTS

We acknowledge support from The National Autonomous University of Mexico (UNAM) and Alianza UCMX of the University of California (UC), through the project included in the Special Call for Binational Collaborative Projects addressing COVID-19. MN is partially supported by CONICET, Argentina. RAB was financially supported by Conacyt through project 283279. We thank Dr. YangQuan Chen for enlightening discussions, to Dr. Daniel Garcia for carefully reading the manuscript and Dr. Florencia Grinbladt for downloading the data.

## Notes

### Competing Interest Statement

The authors have declared no competing interest.

## References

1 D. G. McNeil Jr., “Covid-19: How much herd immunity is enough,” (2020).

2 R. A. Dandekar, S. G. Henderson, M. Jansen, S. Moka, Y. Nazarathy, C. Rackauckas, P. G. Taylor, and A. Vuorinen, medRxiv (2020).

3 C. Rackauckas, Y. Ma, J. Martensen, C. Warner, K. Zubov, R. Supekar, D. Skinner, and A. Ramadhan, arXiv preprint 2001.04385 (2020).

4 R. Dandekar, C. Rackauckas, and G. Barbastathis, Patterns 1, 100145 (2020).

5 E. Alvarez, D. Obando, S. Crespo, E. Garcia, N. Kreplak, and F. Marsico, medRxiv (2020), 10.1101/2020.10.09.20210351, publisher: Cold Spring Har-bor Laboratory Press.

6 R. T. Q. Chen, Y. Rubanova, J. Bet- tencourt, and D. K. Duvenaud, in Advances in Neural Information Processing Systems, Vol. 31, edited by S. Bengio, H. Wallach, H. Larochelle, K. Grauman, N. Cesa-Bianchi, and R. Garnett (Curran Associates, Inc., 2018) pp. 6571–6583.

7 Delphi Group, “Covid symptom survey.” (2020).

8 This definition is in line with the working definition of CLI used by the US Centers for Disease Control and Prevention (CDC) and mirrors the standard definition of influenza-like illness or ILI (defined as fever of at least 100 °F, along with sore throat or cough).

9 Delphi Group, “Covidcast interactive map.” (2020).

10 D. C. Farrow, L. C. Brooks, A. Rumack, R. J. Tibshirani, and R. Rosenfeld, “Delphi epidata api,” (2015).

11 A. Reinhart and R. Tibshirani, “COVID-19 Symptom Surveys through Facebook,” (2020).

12 W. O. Kermack and A. G. McKendrick, Bulletin of Mathematical Biology 53 (1991), https://doi.org/10.1007/BF02464423.

13 D. A. Winkler and T. C. Le, Molecular informatics 36, 1600118 (2017).

14 H. Lin and S. Jegelka, in Advances in Neural Information Processing Systems (2018) pp. 6169–6178.

15 K. He, X. Zhang, S. Ren, and J. Sun, in IEEE Conference on Computer Vision and Pattern Recog https://ieeexplore.ieee.org/document/7780459

16 D. P. Kingma and J. Ba, arXiv preprint 1412.6980 (2014).

17 L. S. Pontryagin, Mathematical Theory of Optimal Processes (CRC Press, 1987) google-Books-ID: kwzq0F4cBVAC.

18 A. Gholami, K. Keutzer, and G. Biros, arXiv preprint 1902.10298 (2019).

19 D. Onken and L. Ruthotto, arXiv preprint 2005.13420 (2020).

20 R. Serban and A. C. Hindmarsh, 1603 (1984). CVODES: An ODE solver with sensitivity analysis capabilities Tech. Rep. (2003).

21 C. Rackauckas, Y. Ma, J. Martensen, C. Warner, K. Zubov, R. Supekar, D. Skinner, A. Ramadhan, and A. Edelman, arXiv preprint 2001.04385 (2020).

22 Dr. Florencia Grinbladt downloaded the data and applied the smoothig.

23 R. T. Alex Reinhart, “Delphi Epidata API,” (2020).

24 C. Rackauckas, et al., “Training a neural ordinary differential equation with mini-batching,” (2020).

25 D. D. Morrison, J. D. Riley, and J. F. Zancanaro, Communications of the ACM 5, 613 (1962). 2itHio.nG(.CBVocPkRa)nd K.-J. Plitt, IFAC Proceedings Volumes 17, (2016) pp. 770–778.

26 H. G. Bock and K.-J. Plitt, IFAC Proceedings Volumes 17, 1603 (1984).

27 H. G. Bock, M. M. Diehl, D. Leineweber, and J. P. SchlÖder, in Nonlinear model predictive control (Springer, 2000) pp. 245–267.

28 C. Tsitouras, Computers & Mathematics with Applications 62, 770–775 (2011).

29 C. Rackauckas and Q. Nie, Journal of Open Research Software 5 (2017).

30 P. Ramachandran, B. Zoph, and Q. V. Le, 1710.05941 [cs] (2017), 1710.05941.

31 The pandas development team, “pandas-dev/pandas: Pandas,” (2020).

32 J. D. Hunter, Computing In Science & Engineering 9, 90 (2007).

33 B. Harrington et al., “Inkscape,” (2004).

34 C. Rackauckas et al., “Diffeqflux: Generalized physics-informed and scientific machine learning (sciml),” (2020).

35 M. Nunñez et al., in preparation (2021).

