## Supplemental Material for "Forecasting virus outbreaks with social media data via neural ordinary differential equations"

Title:

*Matías Nuñez, Nadia L. Barreiro, Rafael A. Barrio and Christopher Rackauckas*

The full survey can be found in the following link: https://cmu-delphi.github.io/delphi-epidata/symptom-survey/coding.html#wave-1

The extracted signals from the survey data are updated daily and can be seen here: https://delphi.cmu.edu/covidcast/survey-results/?date=20210124

An interactive map with all the signals can be seen here https://delphi.cmu.edu/covidcast/

The following is an extract of the survey questions from which the signal where constructed.

### Survey of COVID-Like Illness - US Expansion

Start of Block: Screener

You must be 18 years or older to take this survey. Are you 18 years or older?

- Yes (1)
- No (2)

Start of Block: Section A: Symptoms (forecast)

A1 In the past 24 hours, have **you or anyone in your household** experienced any of the following:

| | Yes (1) | No (2) |
|---|---|---|
| Fever (100°F or higher) (1) | ○ | ○ |
| Sore throat (2) | ○ | ○ |
| Cough (3) | ○ | ○ |
| Shortness of breath (4) | ○ | ○ |
| Difficulty breathing (5) | ○ | ○ |

A2 How many people in your household **(including&nbspyourself)** are **sick** (**fever**, along with **at least one other symptom** from the above list)?

A2b How many people are there in your household **in total (including yourself)**?

A3 What is your current ZIP code?

A4 How many **additional** people in your local community that you know personally are **sick** (**fever**, along with **at least one other symptom** from the above list)?

Start of Block: Section B: Symptoms (non-forecast)

B2 The rest of the survey will go into more detail to get a better understanding of your personal experience.

**In the past 24 hours**, have **you personally** experienced any of the following symptoms? (Select all that apply.)

- ☐ Fever (1)
- ☐ Cough (2)
- ☐ Shortness of breath (3)
- ☐ Difficulty breathing (4)
- ☐ Tiredness or exhaustion (5)
- ☐ Nasal congestion (6)
- ☐ Runny nose (7)
- ☐ Muscle or joint aches (8)
- ☐ Sore throat (9)
- ☐ Persistent pain or pressure in your chest (10)
- ☐ Nausea or vomiting (11)
- ☐ Diarrhea (12)
- ☐ Loss of smell or taste (13)
- ☐ Other (Please specify): (14) ________________________________
- ☒ None of the above (15)

B2b How long, in days, have you been experiencing these symptoms?

B3 You mentioned that you had a fever **in the past 24 hours**. Have you taken your temperature?

- ◯ Yes (1)
- ◯ No (2)

Q40 What was your highest temperature, in °F?

B4 You mentioned that you experienced a cough **in the past 24 hours**. Did you cough up mucus?

- ◯ Yes, I had a lot of mucus (1)
- ◯ Yes, I had a little mucus (2)
- ◯ No, I had a dry cough (3)

B5 Have you been tested for COVID-19 (coronavirus) for your current illness?

- ◯ Yes, I was tested, and received a positive diagnosis for COVID-19 (1)
- ◯ Yes, I was tested, but it was negative for COVID-19 (2)
- ◯ Yes, I was tested, but have not received the result (3)
- ◯ No, I tried to get tested but could not get a test (4)
- ◯ No, I have not tried to get tested (5)

B6 **In the past 24 hours**, have you been to the hospital to seek care for your current illness?

- ◯ Yes (1)
- ◯ No (2)
- ◯ I have tried, but been unable to receive care (3)

C1
Have you ever been told by a doctor, nurse, or other health professional that you have any of the following medical conditions?

(Please select all that apply)

- ☐ Diabetes (1)
- ☐ Cancer (other than skin cancer) (2)
- ☐ Heart disease (3)
- ☐ High blood pressure (4)
- ☐ Asthma (5)
- ☐ Chronic lung disease such as COPD or emphysema (6)
- ☐ Kidney disease (7)
- ☐ Autoimmune disorder such as rheumatoid arthritis or Crohn's disease (8)
- ☐ ⊗None of the above (9)

C2 Have you had a flu shot in the last 12 months?

- ○ Yes (1)
- ○ No (2)

C3 **In the past 5 days**, have you gone to work outside of your home?

- ○ Yes (1)
- ○ No (2)

C4 **In the past 5 days**, have you worked or volunteered in a hospital, medical office, ambulance service, first responder services, or any other health care setting?

- Yes (1)
- No (2)

C5 **In the past 5 days**, have you worked at or visited a long-term care facility or nursing home?

- Yes (1)
- No (2)

C6 **In the past 5 days**, have you traveled outside of your state?

- Yes (1)
- No (2)

C7 To what extent are you intentionally avoiding contact with other people?

- All of the time (1)
- Most of the time; I only leave my home to buy food and other essentials (2)
- Some of the time; I have reduced the amount of times I am in public spaces, social gatherings, or at work (3)
- None of the time (4)

C8 **In the past 5 days**, how often have you ...

| | None of the time (1) | Some of the time (2) | Most of the time (3) | All the time (4) |
|---|---|---|---|---|
| felt nervous, anxious, or on edge? (1) | ○ | ○ | ○ | ○ |
| felt depressed? (2) | ○ | ○ | ○ | ○ |

C9 How do you feel about the possibility that you or someone in your immediate family might become seriously ill from COVID-19 (coronavirus disease)?

- ○ Very worried (1)
- ○ Somewhat worried (2)
- ○ Not too worried (3)
- ○ Not worried at all (4)

C10 **In the past 24 hours**, with how many people have you had direct contact, **outside of your household**? Your best estimate is fine.["Direct contact" means: a conversation lasting more than 5 minutes with a person who is closer than 6 feet away from you, or physical contact like hand-shaking, hugging, or kissing.]

| | Number (1) |
|---|---|
| At work (1) | |
| Shopping for groceries and other essentials (2) | |
| At social gatherings (3) | |
| Other (4) | |

C11 **In the past 24 hours**, have you had direct contact with anyone who recently tested positive for COVID-19 (coronavirus)?["Direct contact" means: a conversation lasting more than 5 minutes with a person who is closer than 6 feet away from you or physical contact like hand-shaking, hugging, or kissing.]

○ Yes (1)

○ Not to my knowledge (2)

C12 Was this person a member of your household?

○ Yes (1)

○ No (2)

A3b In which state are you currently staying?

○ Alabama (1)

○ Alaska (2)

○ Arizona (3)

○ Arkansas (4)

○ California (5)

○ Colorado (6)

○ Connecticut (7)

○ Delaware (8)

○ District of Columbia (9)

○ Florida (10)

○ Georgia (11)

○ Hawaii (12)

○ Idaho (13)

○ Illinois (14)

○ Indiana (15)

○ Iowa (16)

○ Kansas (17)

○ Kentucky (18)

○ Louisiana (19)

○ Maine (20)

○ Maryland (21)

○ Massachusetts (22)

○ Michigan (23)

○ Minnesota (24)

○ Mississippi (25)

○ Missouri (26)

○ Montana (27)

○ Nebraska (28)

○ Nevada (29)

○ New Hampshire (30)

○ New Jersey (31)

○ New Mexico (32)

○ New York (33)

○ North Carolina (34)

○ North Dakota (35)

○ Ohio (36)

○ Oklahoma (37)

○ Oregon (38)

○ Pennsylvania (39)

○ Puerto Rico (40)

○ Rhode Island (41)

○ South Carolina (42)

- South Dakota (43)
- Tennessee (44)
- Texas (45)
- Utah (46)
- Vermont (47)
- Virginia (48)
- Washington (49)
- West Virginia (50)
- Wisconsin (51)
- Wyoming (52)
- I do not reside in the United States (53)

D1 What is your gender?

- Male (1)
- Female (2)
- Non-binary (3)
- Prefer to self-describe: (4) ________________________________________________
- Prefer not to answer (5)

D1b Are you currently pregnant?

- ○ Yes (1)
- ○ No (2)
- ○ Prefer not to answer (3)
- ○ Not applicable (4)

D2 What is your age?

- ○ 18-24 years (1)
- ○ 25-34 years (2)
- ○ 35-44 years (3)
- ○ 45-54 years (4)
- ○ 55-64 years (5)
- ○ 65-74 years (6)
- ○ 75 years or older (7)

D3 How many children **under 18 years old** currently stay in your household?

D4 How many adults **between 18 and 64 years old** currently stay in your household (not including yourself)?

D5 How many adults 65 years old or older currently stay in your household (not including yourself)?

Q36 How much of a threat would you say the coronavirus outbreak is to your household's finances?

- ○ A substantial threat (1)
- ○ A moderate threat (2)
- ○ Not much of a threat (3)
- ○ Not a threat at all (4)
